# Harnessing the power of polygenic risk scores to predict type 2 diabetes and its subtypes in a high-risk population of British Pakistanis and Bangladeshis in a routine healthcare setting

**DOI:** 10.1101/2021.07.12.21259837

**Authors:** Sam Hodgson, Qin Qin Huang, Neneh Sallah, Genes & Health Research Team, Chris J Griffiths, William G Newman, Richard C Trembath, Thomas Lumbers, Karoline Kuchenbaecker, David A. van Heel, Rohini Mathur, Hilary Martin, Sarah Finer

## Abstract

**Background:** Type 2 diabetes is highly prevalent in British Pakistanis and Bangladeshis (BPB). The Genes & Health (G&H) cohort offers means to explore genetic determinants of disease in BPBs.

**Methods:** We assessed whether common genetic loci associated with type 2 diabetes in European-ancestry individuals (EUR) replicate in G&H. We constructed a type 2 diabetes polygenic risk score (PRS) and combined it with a clinical risk instrument (QDiabetes) to build a novel, integrated risk tool (IRT). We compared IRT performance using net reclassification index (NRI) versus QDiabetes alone.We compared PRS distribution between type 2 diabetes subgroups identified by clinical features at diagnosis.

**Findings:** We replicated fewer loci in G&H (n = 76/338, 22%) than would be expected given power if all EUR-ascertained loci were transferable (n = 95, 28%) (p-value = 0.01). In 13,648 patients free from type 2 diabetes followed up for 10 years, NRI was 3.2% for IRT versus QDiabetes (95% confidence interval 2.0 - 4.4%). IRT performed best in reclassification of young adults deemed low risk by QDiabetes as high risk. PRS was independently associated with progression to type 2 diabetes after gestational diabetes (p = 0.028). Mean type 2 diabetes PRS differed between phenotypically-defined type 2 diabetes subgroups (p = 0.002).

**Interpretation:** The type 2 diabetes PRS has broad potential clinical application in BPB, improving identification of type 2 diabetes risk (especially in the young), and characterisation of subgroups at diagnosis.

**Funding:** Wellcome Trust, MRC, NIHR, and others.

**Research in Context:** *Evidence before this study:* People of south Asian origin are disproportionately affected by type 2 diabetes, yet are underrepresented in genetic studies assessing its causation. To date, there have been no published studies that systematically assess how type 2 diabetes genetic risk loci identified in European individuals can be transferred into south Asians, taking into account power and differences in linkage disequilibrium, nor has the clinical utility of a type 2 diabetes polygenic risk score (PRS) been evaluated in this ethnic group. For coronary artery disease, integration of PRS with clinical risk tools has been shown to enhance the prediction of incident disease, in multiple ancestral groups. For type 2 diabetes, whilst it is known from multiple studies of Europeans that PRS can enhance prediction of incident disease, no study has examined PRS performance when integrated with an existing clinical risk tool, although it has potentially significant clinical impact. The identification of type 2 diabetes subgroups at disease presentation has now been studied extensively, but the influence of polygenic risk in characterising these subgroups has not been tested. We examined prior evidence using multiple updated searches across MEDLINE, CINAHL, EMBASE, MEDRXIV and BIORXIV on 29/6/2021 with terms including “type 2 diabetes” and “polygenic risk scor$,” “genetic risk scor$”, “subgroup”, and “cluster” did not identify similar published work.

*Added value of this study:* In the first study to systematically assess the transferability of genetic loci associated with type 2 diabetes in European ancestry individuals into a British Pakistanis and Bangladeshis (BPBs), we find fewer transferable loci than would be expected, accounting for power. We also construct a type 2 diabetes PRS for BPBs and show that its integration with QDiabetes enhances 10-year prediction of incident type 2 diabetes, especially in individuals aged less than 40 years deemed low risk by QDiabetes alone, who tended to be free from comorbidities, and relatively slim. Additionally, we find the PRS is independently associated with progression from gestational diabetes mellitus to type 2 diabetes in BPBs, replicating previous findings in European individuals. We replicate previously-reported subgroups of type 2 diabetes in BPBs, including Mild Age-Related Diabetes, Mild Obesity-Related Diabetes, and Insulin-Resistant Diabetes, and show that PRS distribution differs between clinically-defined clusters. In a novel clustering approach integrating PRS with clinical features, we identify a previously unreported subgroup we name “Clinically Undifferentiated High Polygenic Susceptibility Diabetes”, and observe differences in rates of progression to micro- and macrovascular complications between subgroups.

*Implications of all the available evidence:* A single, low-cost genotyping array can now determine the polygenic risk of multiple diseases in parallel at any point in the life course. We build on existing genomic resources to build a type 2 diabetes PRS that can be used to predict incident disease in a specific ancestral group that is disproportionately affected by the condition. We show that a PRS, when integrated with an established and well-validated clinical risk algorithm, has significant potential clinical utility as both a means to better estimate individual disease risk, and to elucidate the influence of genetics on disease subgroups to aid future efforts to stratify care and treatment of the disease.

## Introduction

Clinical risk prediction of type 2 diabetes is routinely used in healthcare systems to identify at-risk individuals to target preventative care. Contemporary advances in genomic technology, and robust genome-wide association studies have provided a platform with which to develop polygenic risk scores (PRS) to aid the individualised clinical prediction of common complex diseases (1–6). To date, PRS have been developed and tested predominantly in highly selected White European populations with bias towards healthy and older people (7).

There is considerable potential to use PRS to improve the prediction of type 2 diabetes risk (7), particularly when combined with well-validated clinical risk instruments, such as QDiabetes, already in routine use. There is an additional need to study PRS in understudied groups who are at high risk of type 2 diabetes, including people of south Asian ancestry (7,8) and women with a history of gestational diabetes mellitus (GDM) (9). Building enhanced risk tools that combine PRS with tools such as QDiabetes could offer significant opportunities for clinical benefit including enhanced individualised screening and preventive measures such as referral to diabetes prevention programmes (10). PRS may also enhance the characterisation of type 2 diabetes ‘subgroups’, a recent area of significant research and clinical interest due to their potential to valuably capture important heterogeneous features at diabetes diagnosis associated with common biological disease pathways that may predict future diabetes complications and treatment responsiveness (11–13). How and whether polygenic risk of type 2 diabetes is associated with these type 2 diabetes subgroups is currently unclear (14).

In this study, we aimed to develop and evaluate a type 2 diabetes polygenic risk score in British Bangladeshis and Pakistanis (BPBs) enrolled in the Genes & Health (G&H) programme (15). This real-world, community-based cohort (n=48,144) combines genetic data with rich, lifelong electronic health record data and comprises a minority ethnic group living predominantly in socioeconomically deprived circumstances, otherwise underrepresented in health research (15). BPB individuals tend to develop type 2 diabetes at younger ages and at a lower body mass index than European-ancestry individuals(16), but the extent to which genetic aetiology of the conditions is shared between these populations is not well understood. We therefore aimed, firstly, to examine the transferability of type 2 diabetes genetic loci already ascertained in individuals of European ancestry (EUR) to British Pakistani and Bangladeshi individuals (BPBs), and to optimise a PRS for this population. Secondly, we tested whether the PRS enhances the prediction of incident type 2 diabetes when integrated with the commonly-used clinical risk score, QDiabetes (17). Thirdly, we sought to investigate whether the PRS alone might predict the progression to type 2 diabetes from gestational diabetes in BPB women, as has been observed in European and South-East Asian populations (18,19). Finally, we explored whether the PRS might predict clinically heterogeneous type 2 diabetes subgroups at diagnosis. Our findings have major clinical relevance to a wider UK and global population of people of south Asian ancestry at risk of type 2 diabetes.

## Methods

### Study population

Genes & Health (G&H) recruits British Pakistani and British Bangladeshi (BPB) people aged 16 years and above, predominantly in community and primary care settings. We used the February 2020 data release(20) which comprises electronic health record (EHR) data and genotype data from the Illumina Infinium Global Screening Array V3 Chip on 22,944 participants(15). We applied specific inclusion and exclusion criteria to the G&H population for each analysis, described below. Descriptions of quality control and imputation of genotype data are provided in **Supplementary File 1**.

### Ascertainment of conditions using clinical coding

Type 2 diabetes, gestational diabetes mellitus and associated complications were ascertained using clinical codes extracted from primary and secondary care EHR, presented in full in **Table S1**, developed using widely-used clinical coding resources (21). We excluded individuals with clinical codes of conditions causing secondary or monogenic diabetes.

### Transfer of previously-identified GWAS loci to the study population

We obtained previously identified genome-wide association study (GWAS) loci associated with type 2 diabetes in people of European-ancestry (EUR) using Vujkovic *et al*. (22). We performed a GWAS of type 2 diabetes in G&H, and assessed if previously-identified loci were reproducible in G&H at P-value <0.05 (**Supplementary File 1**) using previously-established methods (6). The expected power for replication was estimated, assuming the same effect size as in the EUR discovery sample, and accounting for the allele frequency and sample size in G&H. Even though the same locus and target gene may affect type 2 diabetes risk in both populations, it is possible that this is caused by distinct causal variants in the region. Therefore, trans-ancestry colocalisation analysis was performed using TEColoc to assess whether a transferable locus shared the same causal variant between BPB and EUR populations, using UK Biobank (UKBB) EUR individuals for the latter (23). We applied the Popcorn algorithm to estimate the trans-ancestry genetic correlation between BPB and UKBB EUR populations (24).

### Polygenic risk score construction

We applied previously constructed scores from the PGS Catalog(25), principally developed in EUR populations, to participants in G&H (**see Table S2**). These were compared to PRS optimised within G&H using the clumping and p-value thresholding (C+T) method implemented in PRSice2 v2.2.11 (26,27) based on the largest published type 2 diabetes multi-ancestry GWAS to date(22,25) (**Figure S1)**. For the C+T score, LD estimated from EUR samples (N=503) from the 1000 Genomes project was used for clumping (r^2^=0.1). We calculated multiple scores using various P-value thresholds. We selected the PRS with the highest predictive accuracy in an independent set of samples that were not included in the QDiabetes analysis described below i.e. prevalent cases with onset before the assessment date, and controls who had insufficient data on the assessment date to be included. The area under the receiver operating characteristic curve (AUC) was estimated with the R package “pROC”. Predictive accuracy of PRS was quantified by incremental AUC, or the gain in AUC when adding PRS to the reference model which accounted for participant age, gender, and 10 genetic principal components (PCs). The C+T PRS showed higher predictive accuracy than scores from the PGS Catalog, and was thus used in downstream analysis (**Figure S1**). A scaled PRS following the normal distribution with mean and median of 0 and standard deviation of 1 was constructed across BPB participants after regressing out the first ten PCs, allowing direct comparison between ancestry groups(28).

### Development of an integrated risk tool to predict incident type 2 diabetes

We assessed whether PRS can enhance 10-year risk prediction of type 2 diabetes compared to QDiabetes, a validated, EHR-based risk prediction tool commonly used in UK primary care to estimate an individual’s 10-year risk of developing type 2 diabetes(17). There are three QDiabetes models. Model A provides estimates based on age, ethnicity, family history, comorbidities, and prescribed medications. Model B (which has the highest clinical predictive value) uses the same variables as model A, plus fasting plasma glucose (FPG), while model C is composed of model A plus HbA1c. Analysis was performed in 13,648 individuals aged 25-84 years with no prior history of type 2 diabetes as of 1st May 2010, no recorded value of HbA1c > 48 mmol / mol or FPG > 7.0mmol/L (**Table S1**). These inclusion criteria aligned with those used for development and validation of the QDiabetes model (17). QDiabetes scores were calculated on the assessment date of 1st May 2010 for each participant (see **Table 1** for participant characteristics) on the basis of available clinical data required to run the model. For each individual, the QDiabetes risk estimate was combined with the PRS to form an integrated estimate of risk of developing type 2 diabetes over the next 10 years (Integrated Risk Tool, IRT), as previously described (1). 10-year risk of disease was classified as high (>10%) or low (<10%) in line with (1). Relative model performance was assessed using the concordance index (C-Index) and net reclassification index (NRI; the proportion of participants correctly reclassified as high or low risk by the IRT compared to QDiabetes, minus the proportion of participants incorrectly reclassified) in all individuals and in age-by-sex subgroups (1,7). In a separate analysis restricted to women with a history of GDM, we compared characteristics (including type 2 diabetes PRS) of those who subsequently did (n = 127) and did not (n = 175) develop type 2 diabetes. Further details of incident disease prediction are provided in **Supplementary File 1**.

**Table 1:** Characteristics of participants included in analysis with each QDiabetes model. QDiabetes Model A is calculated in individuals without HbA1c or fasting plasma glucose (FPG); Model B by adding FPG to Model A; and Model C by adding HbA1c to Model A. Values show the mean with standard deviation in brackets, unless otherwise indicated. Index of Multiple deprivation presents Means were compared with analysis of variance (ANOVA).

|  | **Model A (N = 13648)** | **Model B (N=4334)** | **Model C (N=864)** | **p value** |
| --- | --- | --- | --- | --- |
| % Developing type 2 diabetes | 18.2 | 27.0 | 33.3 | < 0.001 |
| Type 2 Diabetes Polygenic Risk Score | -0.046 (0.99) | -0.080 (0.99) | -0.062 (1.01) | 0.210 |
| Age (Years) | 35.4 (8.8) | 42.0 (11.1) | 45.2 (12.3) | < 0.001 |
| % Female | 49.8 | 59.2 | 64.9 | < 0.001 |
| % Bangladeshi | 69.9 | 73.9 | 58.0 | < 0.001 |
| % Family History of Diabetes | 35.9 | 40.2 | 33.9 | <0.001 |
| Index of Multiple Deprivation (2015) score | 7.4 (2.2) | 7.8 (1.9) | 7.2 (2.4) | < 0.001 |
| BMI | 25.8 (4.6) | 26.704 (4.624) | 27.7 (4.7) | < 0.001 |
| HbA1c | 39.1 (4.3) | NA | 38.0 (4.2) | 0.271 |
| Fasting Glucose | 4.9 (0.7) | 4.8 (0.7) | NA | 0.154 |
| HDL | 1.2 (0.3) | 1.2 (0.3) | 1.2 (0.3) | < 0.001 |
| Triglycerides | 1.9 (1.3) | 1.8 (1.2) | 1.7 (1.2) | 0.077 |
| **Pre-existing Conditions** |  |  |  |  |
| Gestational diabetes (% females) | 4.1 | 5.2 | 10.7 | < 0.001 |
| Polycystic ovarian syndrome (% females) | 4.7 | 7.3 | 6.6 | < 0.001 |
| Cardiovascular disease (%) | 2.5 | 4.7 | 9.7 | < 0.001 |
| Hypertension (%) | 6.1 | 14.2 | 13.7 | < 0.001 |

### Identification of type 2 diabetes subgroups and their association with PRS

We applied data-driven clustering techniques to define subgroups of individuals with distinct characteristics in two separate models using latent class analysis (Stata V16.0) based on methodology used by (11–13). The optimal number of clusters was selected on the basis of information criteria elbow plots (**Figure S2)** and was found to be five for all model iterations. The first model (Clinical Model) was based on five clinical variables at the time of type 2 diabetes diagnosis: age, body mass index (BMI), HbA1c, serum triglycerides and serum high-density lipoprotein (HDL). Analysis was restricted to 4,266 participants for whom values for these variables were available within 1 year of type 2 diabetes diagnosis. Differences in PRS between clusters were compared using analysis of variance (ANOVA) (RStudio V1.1.413). In the second model (Integrated Model) we repeated clustering using age, BMI, HbA1c, serum triglycerides and PRS as covariates in 5,905 individuals with available data within 12 months of diagnosis. HDL was omitted from the Integrated Model as its inclusion did not alter cluster characteristics, but did reduce sample size. For both models, progression to micro- and macrovascular complications (defined in **Table S1)** was compared between clusters with Cox proportional hazard models controlling for hypertension, statin use, and serum total cholesterol using the R package “survival” (V2.3-11) in individuals free from micro- and macrovascular complications at the time of type 2 diabetes diagnosis.

## Results

As of May 1st 2020 (the end of the study period), 7,599 individuals with an EHR diagnosis of type 2 diabetes were enrolled in G&H, followed up for a mean of 9.69 years after diagnosis. 52.8% of these individuals were in the most deprived Index of Multiple Deprivation quintile in the UK. Mean age at time of diagnosis was 46.2 years. 1205 individuals (15.9%) developed macrovascular complications between type 2 diabetes diagnosis and the end of the study period; 2300 (30.3%) developed microvascular complications.

### Genetics of type 2 diabetes in BPB versus European individuals

We first investigated the extent to which genetic risk for type 2 diabetes was shared between G&H (BPB - British Pakistani and Bangladeshis) and EUR individuals. Results are described in more detail in **Supplementary File 1**. In brief, we found that trans-ethnic genetic correlation (i.e. the correlation of causal-variant effect sizes) between G&H and UKBB EUR individuals was significantly lower than 1 (r_g_ = 0.68, standard error = 0.15, p-value (for the null hypothesis that r_g_ = 1) = 0.03). Most of the 338 genetic loci ascertained in EUR populations which were well-powered to replicate in G&H did so (76 (22.5%) replicated versus 28.1% expected; binomial p=0.01; **Table S3**). Of 27 replicated loci assessed, 9 (33%) had significant evidence of shared causal variants (**Table S4**). For example, we observed shared causal variants at the *TCF7L2* locus, one of the known loci with the strongest association with type 2 diabetes, and at the *KCNJ11* locus, which is the target gene for drugs such as Glyburide (29,30).

### Performance of an integrated risk tool to predict incident type 2 diabetes

We constructed a PRS for type 2 diabetes using the C+T method, based on multi-ancestry GWAS meta-analysis (22). This had an odds ratio (OR) per standard deviation (SD) of 1.57 (95% confidence interval [CI]: 1.50–1.65) and an incremental AUC of 0.032 (95% CI: 0.026, 0.039) in G&H. There was no correlation between PRS and QDiabetes scores (Pearson’s coefficients -0.03, 0.08, 0.13 for QDiabetes models A, B, and C respectively; associated p-values 0.31, 0.18, 0.16). Compared to QDiabetes alone, the integrated risk tool (IRT) combining QDiabetes model A (multiple clinical risk factors) with PRS improved the 10-year prediction of type 2 diabetes as assessed by net reclassification index (NRI); NRI 3.22% (95% CI: 2.00 - 4.38%) in 13,648 included individuals (**Figure 1A)**. The IRT C-index, a goodness-of-fit metric approximating the area under the receiver-operator curve, was superior to QDiabetes model A for participants aged less than 40 years (p = 0.002) (**Figure 2A / Table S5)**, but not for people aged over 40 years. Enhancement of type 2 diabetes prediction with the IRT persisted but was attenuated when QDiabetes model B (model A plus fasting plasma glucose) was used: NRI 0.80% (95% CI 0.21 - 1.42%). The IRT did not improve on the QDiabetes score with model C (model A plus HbA1c) (NRI 0.20%; 95% CI: -0.09 - 0.44%) (**Figure 1B and 1C)**. There were no observed differences in C Index between QDiabetes and IRT in models B and C (**Figures 2A and 2B / Table S5)**.

**Figure 1:**
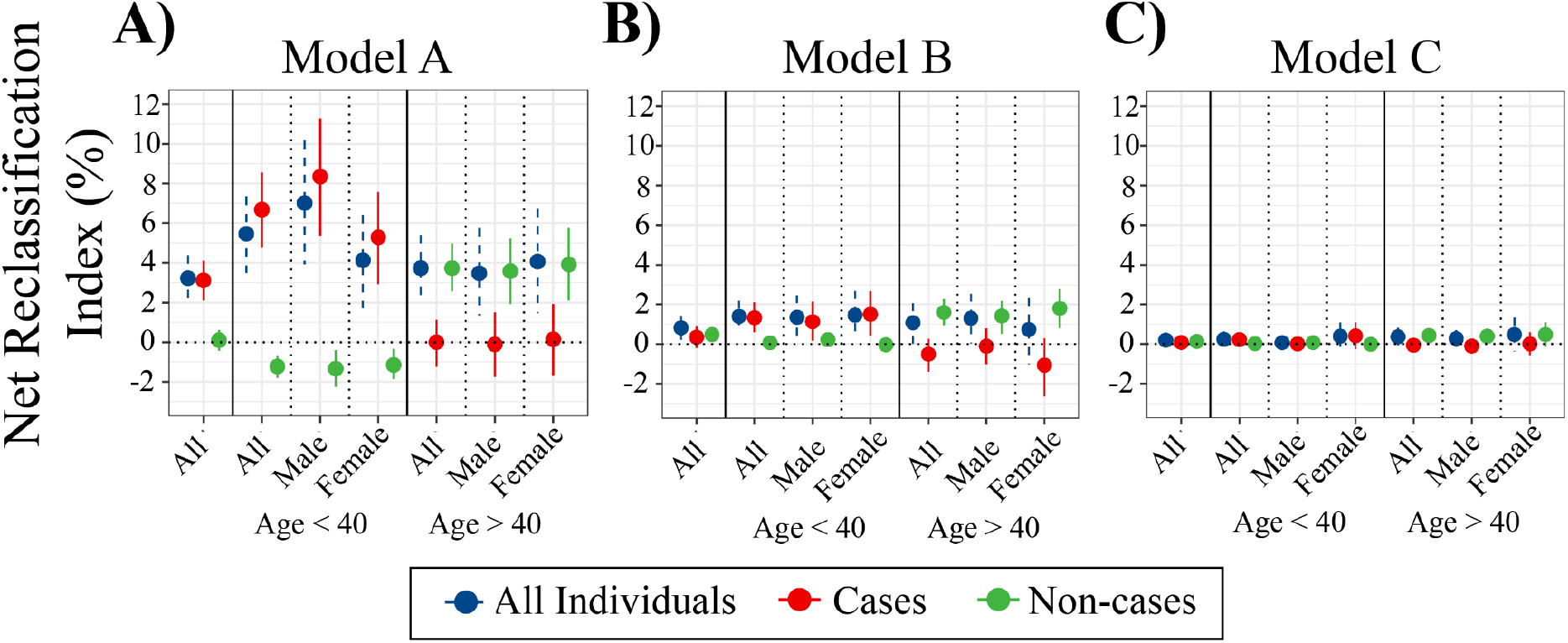
Net Reclassification Index (NRI) and 95% confidence intervals comparing 10 year prediction of type 2 diabetes between QDiabetes models A (n = 13,648), B (n = 4,334), and C (n = 864) and integrated risk tools combining each QDiabetes model with type 2 diabetes polygenic risk score. NRI is presented overall and for age-by-sex subgroups. The NRI for individuals who subsequently develop type 2 diabetes (cases) and who do not develop type 2 diabetes (non-cases) are presented alongside the NRI for cases and non-cases combined (all individuals.

**Figure 2:**
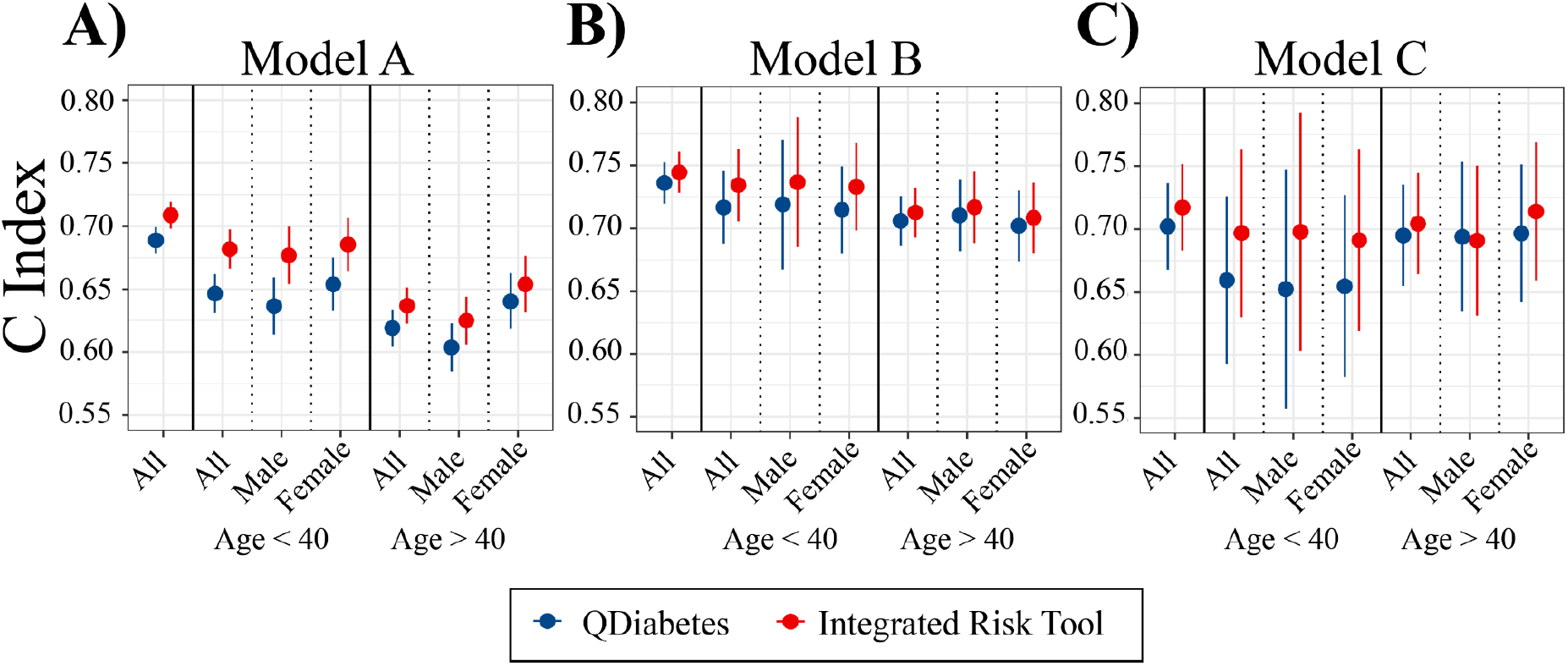
Comparison of C Index calculated from Cox proportional hazard models for QDiabetes models A, B, and C and integrated risk tools combining each QDiabetes model with individual type 2 diabetes polygenic risk score. C index is presented for all participants, and for age-by-sex subgroups.

Across all IRT models (for QDiabetes model A, B and C), NRI was higher in participants aged less than 40 years who subsequently developed type 2 diabetes (cases) than those who did not (non-cases), implying improved ability to correctly classify younger individuals as high risk. The converse pattern was seen in individuals aged greater than 40 years, implying enhanced ability to correctly classify older individuals as low risk. The clinical features of individuals whose risk was reclassified are presented in **Table S6**. The number of individuals reclassified in each model are presented by age band and sex in **Table S7**.

### Prediction of type 2 diabetes after gestational diabetes using PRS

In women who had a history of gestational diabetes (GDM), Type 2 diabetes PRS was higher in those who had subsequently developed type 2 diabetes, compared to those who had not (mean value (SD) 0.408 (0.93) vs 0.140 (0.98), p = 0.02) **(Figure 3)**. Among individuals without type 2 diabetes (non-cases), women with a history of GDM displayed a higher mean PRS than age- and BMI-matched females without GDM, as well as males (p = 0.003 and p=0.001, respectively) (**Figure 3)**. This could be because these individuals have increased propensity to develop type 2 diabetes in the future, but have not yet developed it, or because they have undiagnosed type 2 diabetes, for example, through missed screening tests post-delivery. In multivariate survival analysis restricted to women with a history of GDM, type 2 diabetes PRS was independently associated with development of type 2 diabetes after adjustment for (1) QDiabetes score model A (hazard ratio per SD of PRS (HR) 1.23, 95% CI 1.05 - 1.42; p = 0.028;) and (2) established risk factors for type 2 diabetes (HR 1.37, 95% CI 1.17 - 1.57, p = 0.002) (**Table S8)**. Similar findings were observed in age- and BMI-matched male and female (without a history of GDM) controls (**Table S8)**, indicating that the utility of the type 2 diabetes PRS for risk prediction does not appear to differ between women with versus without GDM.

**Figure 3:**
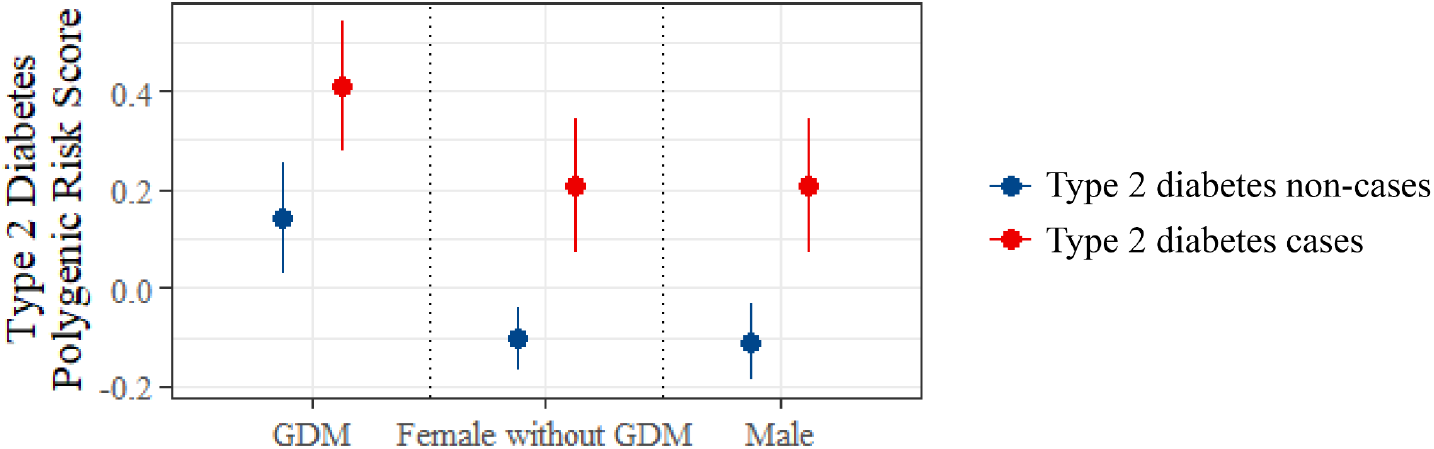
Mean type 2 diabetes polygenic risk score in women with gestational diabetes mellitus (GDM) who subsequently did and did not develop type 2 diabetes, compared to age- and BMI-matched control groups of females without GDM, and males. Data are presented as group mean with 95% confidence intervals according to sex-by-GDM subgroup, stratified by subsequent type 2 diabetes status.

### Type 2 diabetes subgroup identification using clustering

In our Clinical Model, we identified type 2 diabetes subgroups by clustering age, BMI, HBA1c, HDL and triglycerides at the time of diagnosis (**Figure 4A/Table 2**). We replicated some previously identified clusters: Mild Obesity-related Diabetes (MOD), Mild Age-Related Diabetes (MARD), Severe Insulin Resistant Diabetes (SIRD) and Mild Diabetes (MD), and use their previous nomenclature (11–13,31). Our MD cluster may include individuals who are insulin deficient as it is characterised by low serum triglycerides (1.21 (0.43) mmol/L) in addition to high HDL (2.07 (0.25) mmol/L), but in the absence of clinical measures of insulin secretion such as C-peptide, this cannot be confirmed. Our cluster with the largest membership was undifferentiated by clinical features; we therefore call it Clinically-Undifferentiated Diabetes (CUD). There were no statistically significant differences in rates of progression to micro- or macrovascular complications between clusters in the Clinical model. However, we found that the PRS differed significantly between clusters(ANOVA p=0.002) and that CUD was characterised by the highest observed PRS (0.34 (0.97)). PRS was lowest in MOD and MARD (mean PRS (0.07 (1.02) and 0.23 (0.97)), respectively.

**Figure 4:**
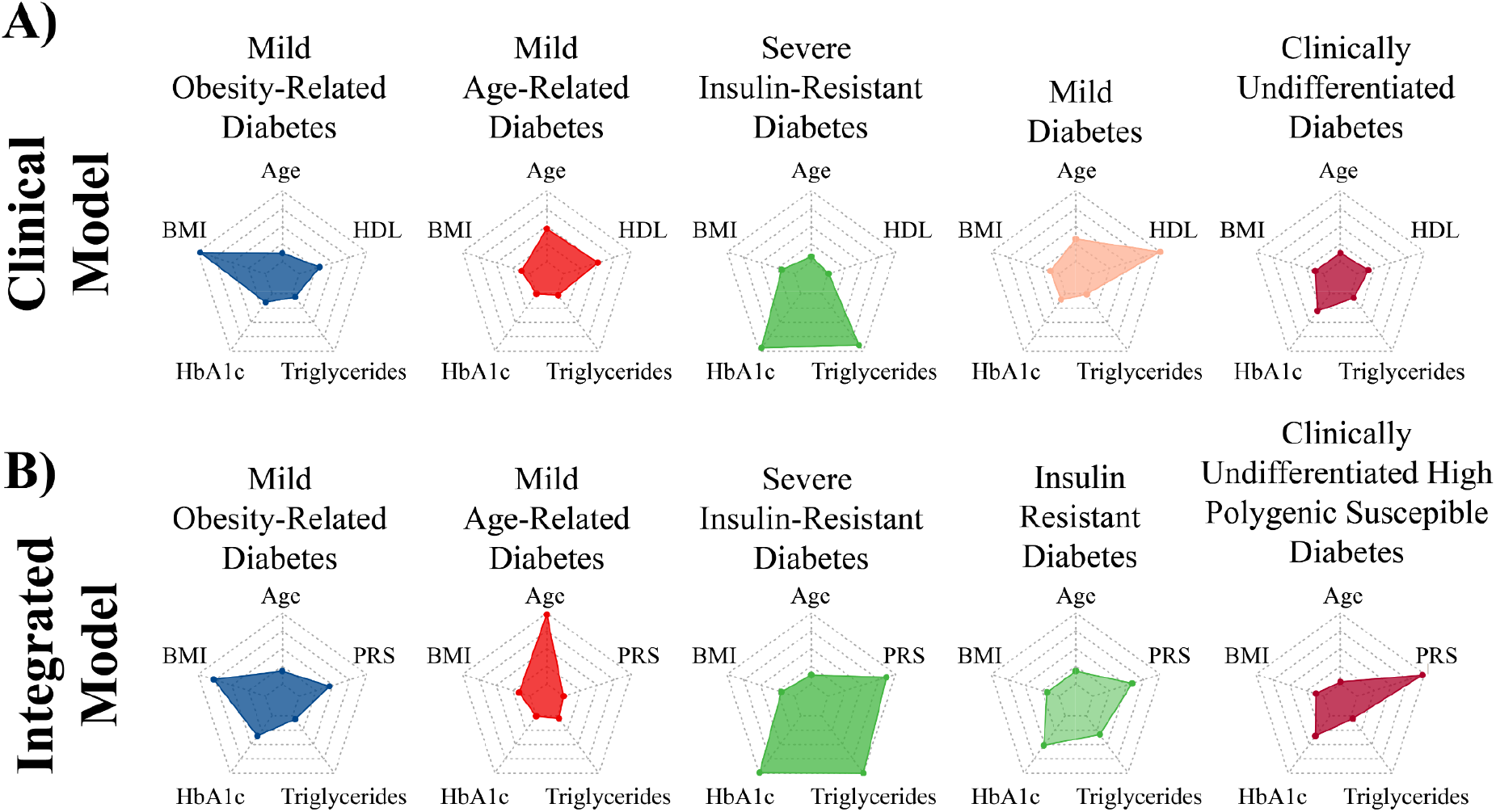
Radar plots showing cluster mean valuess of the variables used to derive five clusters of individuals with type 2 diabetes at the time of diagnosis in two separate models. In panel A (Clinical Model), latent class analysis was performed using age, BMI (body mass index), HbA1c, serum triglycerides, and high density lipoprotein (HDL). In panel B (integrated model), clustering was repeated using type 2 diabetes polygenic risk score (PRS) in place of HDL. The centre and edge of each polygon represents the minimum and maximum mean cluster values for each variable, respectively. All polygons represent the same scale; all scales are linear. Mean values for each variable within each cluster are represented by coloured dots. For example, mean BMI is highest in cluster 1.

**Table 2:** Distribution of clustering variables and type 2 diabetes polygenic risk score (PRS) across five data-driven clusters of individuals with type 2 diabetes at the time of diagnosis in a clinical (panel A) and integrated clinical and genetic (panel B) model. PRS was not used as a clustering variable in the clinical model; it was used as a clustering variable in the integrated model. In the clinical model, the distribution of PRS between groups was compared after cluster allocation. Data are presented as mean (standard deviation). Mean values were compared using ANOVA.

| **A** | **Clinical Model** |  | **Mild Obesity-Related Diabetes (MOD) (N=211)** | **Mild Age-Related Diabetes (MARD) (N=861)** | **Severe Insulin Resistant Diabetes (SIRD) (N=32)** | **Mild Diabetes (MD) (N=95)** | **Clinically Undifferentiated Diabetes (CUD) (N=3067)** | **p value** |
| --- | --- | --- | --- | --- | --- | --- | --- | --- |
|  |  | **Age (Years)** | 44.5 (11.4) | 50.9 (12.2) | 43.1 (10.9) | 49.3 (12.3) | 45.2 (10.6) | **<0.001** |
|  |  | **BMI (kg/m<sup>2</sup>)** | 40.2 (3.6) | 27.6 (4.0) | 28.2 (3.9) | 27.1 (5.2) | 27.5 (3.7) | **<0.001** |
|  |  | **HbA1c (mmol/mol)** | 57.5 (15.8) | 53.4 (13.4) | 71.9 (26.9) | 55.9 (17.63) | 59.0 (16.4) | **<0.001** |
|  |  | **Serum Triglycerides (mmol/L)** | 1.72 (0.73) | 1.39 (0.63) | 12.2 (3.95) | 1.21 (0.43) | 1.97 (1.08) | **<0.001** |
|  |  | **HDL (mmol/L)** | 1.17 (0.24) | 1.45 (0.15) | 0.79 (0.21) | 2.07 (0.20) | 0.99 (0.17) | **<0.001** |
|  |  | **PRS** | 0.07 (1.02) | 0.23 (0.97) | 0.29 (1.14) | 0.26 (1.09) | 0.34 (0.97) | **0.002** |
| **B** | **Integrated Model** |  | **Mild Obesity-Related Diabetes (MOD) (N=556)** | **Mild Age-Related Diabetes (MARD) (N=1180)** | **Severe Insulin-Resistant Diabetes (SIRD) (N=37)** | **Insulin Resistant Diabetes (IRD) (N=579)** | **Clinically Undifferentiated High Polygenic Susceptibility Diabetes (CUPS) (N=3489)** | **P value** |
|  |  | **Age (Years)** | 44.8 (9.6) | 61.6 (7.6) | 43.7 (11.3) | 42.8 (9.9) | 41.6 (7.6) | **<0.001** |
|  |  | **BMI (kg/m<sup>2</sup>)** | 37.2 (3.9) | 27.5 (3.4) | 28.1 (3.7) | 27.8 (3.8) | 26.7 (3.3) | **<0.001** |
|  |  | **HbA1c (mmol/mol)** | 60.0 (17.1) | 53.1 (11.6) | 72.8 (26.8) | 63.3 (19.1) | 60.0 (16.5) | **<0.001** |
|  |  | **Serum Triglycerides (mmol/L)** | 1.77 (0.73) | 1.62 (0.70) | 13.0 (3.63) | 4.97 (1.05) | 1.66 (0.74) | **<0.001** |
|  |  | **PRS** | 0.16 (0.91) | -0.10 (0.92) | 0.41 (0.96) | 0.25 (1.02) | 0.48 (0.95) | **<0.001** |

Given these results, we next explored whether inclusion of PRS as a covariate in the clustering model could further refine the subgroups (henceforth termed “Integrated model”). We included PRS in the latent class analysis along with age, BMI, HbA1c, and triglycerides (**Figure 4B/Table 3)**. We identified broadly similar clusters, although with some additional differentiation not seen in the Clinical Model: MOD and MARD were maintained, but in the latter, mean age was notably higher than in the Clinical Model (61.6 (7.6) versus 50.9 (12.2) years). In this analysis, two clusters representing insulin resistant diabetes were identified, differentiated by both the severity of hypertriglyceridaemia and PRS, and therefore we call these SIRD and Insulin Resistant Diabetes (IRD). The MD cluster was not identified in this analysis. We again identified a prevalent clinically-undifferentiated cluster that was associated with a high PRS (mean PRS 0.48 (0.95)), and we therefore rename this cluster Clinically-Undifferentiated High Polygenic Susceptibility Diabetes (CUPS). In Cox proportional hazard models (**Figure S3**), macrovascular complication rates were higher in MARD and MOD (p<0.001 and p=0.009, respectively); microvascular rates were highest in IRD and MARD (p<0.001 and p=0.002).

## Discussion

In this study, we harness the power of a large population-based study of British Bangladeshi and Pakistani people with linked health and genomic data to improve the prediction of type 2 diabetes. For the first time in a real-world study population, we show that use of PRS enhances prediction of incident type 2 diabetes on top of an established and clinically validated risk prediction tool, QDiabetes. We also show that among BPB women with a history of GDM, PRS is associated with type 2 diabetes development. Additionally, we show that our PRS is variably associated with type 2 diabetes subgroups and can itself distinguish a subgroup that is undifferentiated by clinical features.

This is the first study to systematically assess the transferability of genetic loci associated with type 2 diabetes in EUR individuals into BPB individuals, which is important to understand the extent to which the aetiology of the disease varies, particularly where pathophysiological differences between populations are well-known(13,32). Previous studies have assessed directional consistency of genetic effects(33) or heterogeneity in allelic odds ratio(34) across ancestry groups, but have not explicitly asked how many genetic loci one might expect to replicate in a sample of a given ancestry given the power and linkage disequilibrium differences. We observed a lower percentage of previously identified GWAS loci that were reproducible (22.5%) in G&H than expected (28.1%) after accounting for power, consistent with an only moderate trans-ethnic genetic correlation between the two populations. Among the genomic loci which did transfer between EUR and BPB individuals and were powered to interrogate this, only 33% showed evidence of shared causal variants. Hence, despite the considerable overlap in genetic risk between BPB and EUR individuals, our results should still motivate larger studies of type 2 diabetes genetics in different ancestry groups(35) in order to better characterise the genetic contribution to disease and ancestry-specific aetiological features. In the future, the findings of such studies may point to different effects of risk factors via Mendelian randomisation, or different potential drug targets across populations.

Our integrated risk tool for type 2 diabetes that combines QDiabetes scores with PRS and find that it improved risk classification in a population at high risk of type 2 diabetes. Specifically, the integrated risk tool improved net reclassification index (NRI), compared to QDiabetes models A (3.22%) and B (0.80%). The integrated risk tool did not improve NRI compared to model C, which may reflect reduced power in model C due to lower sample size (n =864) than the other two models (n=13648 for model A; n=4344 for model B). Alternatively, the lower NRI for models B and C could imply that the benefit of integrating genetic information with risk models already incorporating metrics of hyperglycaemia is more limited than models without hyperglycaemic metrics (Model A). Hippisley-Cox and Coupland (17) describe model B (incorporating fasting plasma glucose) as the best-fitting model in 11.5 million (multi-ethnic) individuals in the QResearch database (C index 0.89 for women, 0.87 for men for model B, compared to 0.83 for women and 0.81 for men in model A), but we observed lower C index values for QDiabetes alone in our study (**Figure 2**). The lower C index values for comparable models in our study could be due to differing age and deprivation distributions between the QResearch database and G&H participants, and the lack of ethnic diversity in our model, since ethnicity is strongly weighted in the QDiabetes risk estimation algorithms and explains a large proportion of variance in type 2 diabetes risk.

We observed positive NRIs with the IRT driven principally by enhanced reclassification of younger individuals from low to high risk, and improved downgrading of older individuals from high to low risk. In the context of a growing burden of type 2 diabetes in increasingly resource-constrained health systems, such reclassification would support more effective resource allocation, with intensification of preventative care pathways for those at highest risk (e.g. early referral to type 2 diabetes prevention programmes(10)) and relaxation of those who are identified to be at reduced risk. We find that individuals being reclassified as higher risk tended to be young (mean age 36.0 years), free from comorbidities, and relatively slim (mean BMI 25.9 kg/m^2) **(Table 2)**. According to estimates by the International Diabetes Federation, half of cases of type 2 diabetes remain undiagnosed(36). Identification these individuals as high risk using our IRT, who would likely otherwise have been considered healthy, could offer significant opportunities to identify and manage type 2 diabetes early and prevent subsequent morbidity and mortality (37). This finding is particularly important given the observation that early-onset type 2 diabetes is associated with rapid progression to vascular complications (38).

We show that in British-Pakistani and -Bangladeshi women with a history of GDM, a type 2 diabetes PRS is associated with development of type 2 diabetes. This finding persists after adjustment for established clinical risk factors and the QDiabetes risk score. Our findings are in keeping with prior reports of an association between a type 2 diabetes-PRS and development of type 2 diabetes in white (18) and South-East Asian (19) women with a history of GDM. While previous studies have applied alternative type 2 diabetes-PRSs to women of South Asian origin to identify those at higher risk of GDM(39), none to date have explored their association with type 2 diabetes development in women with GDM. Although we observed similar associations between PRS and type 2 diabetes development in age- and BMI-matched male and female controls, the clinical utility of this finding in women with a history of GDM is clearer. Identifying individuals at higher risk of developing type 2 diabetes after GDM. This finding has significant clinical relevance, to a patient group at extremely high risk of type 2 diabetes, where improvements to follow-up and screening processes achieved by individualisation could be valuable. This is particularly clinicially important in BPB women given their increased risk of developing type 2 diabetes relative to other ethnic groups, and the globally low uptake of postpartum diabetes screening in non-White individuals (40).

We used data-driven clustering approaches in two separate models to explore how PRS might associate with the type 2 diabetes subgroups that are increasingly recognised as a route to developing stratified diabetes care. Despite the absence of biomarkers such as auto-antibodies and C-peptide, which are seldom measured in the routine datasets we had available, we were able to reproduce previously described diabetes subgroups(11–13) in a previously uninvestigated population of BPBs. We show heterogeneous distribution of a type 2 diabetes PRS across clusters in a model whose membership was defined by clinical measures. PRS was lowest in the Mild Age-Related Diabetes cluster, concordant with our findings that it performs best in predicting the risk of type 2 diabetes in people aged under 40 years, and the Mild Obesity-related Diabetes cluster suggesting that other polygenic influences (e.g. on body weight) may be more important. Our Severe Insulin Resistant Diabetes cluster was characterised by markedly raised serum triglycerides and is worthy of further exploration in studies of rare genetic variants. When we incorporated the PRS in an integrated cluster model with clinical features, we observed more distinct classification of subgroups than our previous model, emergence of clear clusters representing Insulin Resistant and Severe Insulin Resistant Diabetes, and clear emergence of a large cluster predicted by high polygenic susceptibility alone, without defining clinical features. This exploratory analysis showed that the diabetes subgroups identified using the integrated cluster model were associated with differential rates of progression to complications which were not apparent in the clusters found by the Clinical Model, implying that addition of PRS to data-driven identification of type 2 diabetes subgroups could provide an unexplored clinical tool to risk-stratify patients effectively target care. Further work, in larger studies and across other ancestry groups, will be needed to validate these findings and understand their biological and clinical relevance.

### Strengths and limitations

Strengths of this study include its assessment of an ethnic minority population with high levels of type 2 diabetes and associated morbidity and mortality that has been under-represented in research studies to date; its use of high quality longitudinal data from a relatively unselected sample and linkage to genetic data. Our genetic analysis is robust and we build a strong justification for the construction and evaluation of ancestry-specific PRSs. The main weaknesses of our work include our need to impute missing data not present in health records and the lack of a replication cohort to externally validate findings. Our analysis of the progression to type 2 diabetes after GDM was limited by the low uptake of postpartum diabetes screening that may have resulted in underdiagnosis of type 2 diabetes and ascertainment bias. Across all of our analyses, it is likely that some individuals coded as having type 2 diabetes actually have type 1 diabetes or rare monogenic forms of diabetes, although the absolute number of these and impact on overall findings is likely to be very small.

### Conclusions

We highlight the need for ancestry-specific genetic tools to be developed for diverse populations to elucidate aetiological differences. Our PRS, optimised in British South Asians, enhances prediction of incident type 2 diabetes when combined with an established clinical risk tool, compared to using the clinical tool alone, and particularly in young people. Additionally, the PRS has value in predicting the onset of type 2 diabetes in a specific high risk group, women affected by gestational diabetes. These findings could aid the personalisation of care of people at risk of type 2 diabetes. The PRS also adds a unique dimension to the identification of diabetes subgroups at diagnosis, and their association with future complications, that may be valuable for stratification of care in the future. Our work provides important insight into the genetic risk in an ethnic group underrepresented in research but disproportionately affected by type 2 diabetes, and has significant potential to be translated into clinical practice.

## Data Availability

Genes and Health aims to provide wide access to a broad range of researchers. Summary data is available online; sequencing and individual-level phenotypic data is available via application. Further details can be found in the following link.

https://www.genesandhealth.org/research/scientists-using-genes-health-scientific-research

## Acknowledgements

Genes & Health is/has recently been core-funded by Wellcome (WT102627, WT210561), the Medical Research Council (UK) (M009017), Higher Education Funding Council for England Catalyst, Barts Charity (845/1796), Health Data Research UK (for London substantive site), and research delivery support from the NHS National Institute for Health Research Clinical Research Network (North Thames). HCM and QQH are funded by the Wellcome Trust Grant 206194 to the Wellcome Sanger Institute. R.T.L is supported, in part, by the BigData@Heart Consortium funded by the Innovative Medicines Initiative-2 Joint Undertaking under grant agreement No. 116074, and the National Institute for Health Research University College London Hospitals Biomedical Research Centre. We thank Social Action for Health, Centre of The Cell, members of our Community Advisory Group, and staff who have recruited and collected data from volunteers. We thank the NIHR National Biosample Centre (UK Biocentre), the Social Genetic & Developmental Psychiatry Centre (King’s College London), Wellcome Sanger Institute, and Broad Institute for sample processing, genotyping.. We thank Barts Health NHS Trust, NHS Clinical Commissioning Groups (Hackney, Waltham Forest, Tower Hamlets, Newham), East London NHS Foundation Trust, Bradford Teaching Hospitals NHS Foundation Trust for GDPR-compliant data sharing. Most of all we thank all of the volunteers participating in Genes & Health.

The Genes and Health Research team includes (in alphabetical order): Shaheen Akhtar, Mohammad Anwar, Elena Arciero, Samina Ashraf, Gerome Breen, Raymond Chung, Charles J Curtis, Maharun Chowdhury, Grainne Colligan, Panos Deloukas, Ceri Durham, Sarah Finer, Chris Griffiths, Qin Qin Huang, Matt Hurles, Karen A Hunt, Shapna Hussain, Kamrul Islam, Ahsan Khan, Amara Khan, Cath Lavery, Sang Hyuck Lee, Robin Lerner, Daniel MacArthur, Bev MacLaughlin, Hilary Martin, Dan Mason, Shefa Miah, Bill Newman, Nishat Safa, Farah Tahmasebi, Richard C Trembath, Bhavi Trivedi, David A van Heel, and John Wright.

## Contributions statement

SF and HM conceived the research question. SF, HM, SH, QQH, RM, KK and TL designed the study. SF, CG, WN, RT, DvH, HM, Genes & Health Research Team lead and conduct the Genes & Health programme, which is the primary data source for this work. SH, QQH, NS performed the statistical analysis, and all authors were involved in the interpretation of results. SH drafted the initial version of the manuscript, and all co-authors contributed intellectually to revisions of the manuscript and the decision to submit for publication

