## Supplementary Figures 1-3 for "Harnessing the power of polygenic risk scores to predict type 2 diabetes and its subtypes in a high-risk population of British Pakistanis and Bangladeshis in a routine healthcare setting"

<u>Figure S1:</u> PRSs constructed using the clumping and P-value thresholding (C+T) method or from the PGS Catalog

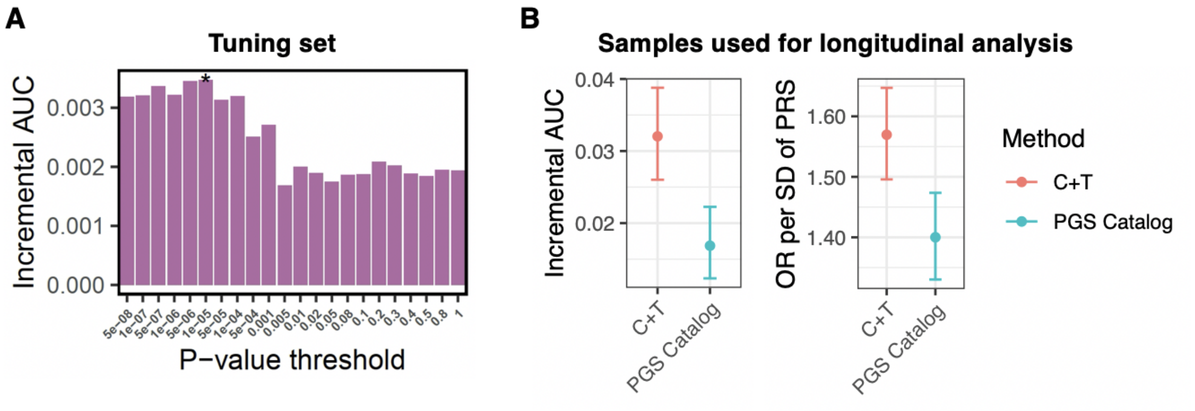


**A**. Incremental AUC of C+T PRSs across various P-value thresholds in the tuning set containing samples that were not included in the main analysis (2,249 cases and 4,443 controls; unrelated). Asterisk indicates the reported PRS with the highest predictive accuracy, which was used in subsequent analyses. **B**. Performance of the C+T PRS (red) and a previously developed score from the PGS Catalog (blue) in the samples used for longitudinal analysis (2,688 cases and 9,221 controls; unrelated). Incremental AUC and odds ratio (OR) per standard deviation (SD) of PRS are compared. Error bars represent 95% confidence intervals. The most accurate score from the PGS Catalog (PGS ID: PGS000020) was developed by Läll K *et al.*[1] using GWAS summary data from the DIAGRAM 2012 study [2].

Figure S2: Elbow plot to identify optimal number of clusters in latent class analysis using Bayesian (BIC) and Akaike (AIC) information criteria. AIC and BIC are penalised likelihood criteria which estimate the difference between the fitted likelihood function of a model, and the estimated true likelihood function of the data set. Lower values for AIC and BIC are associated with better model fit. For all model iterations the optimal number of clusters selected was 5.

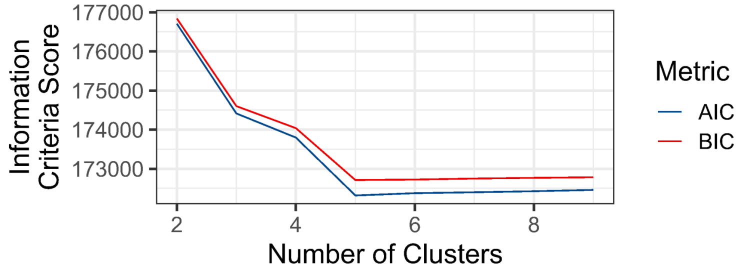

Figure S3: Cox proportional hazard models to show association between cluster membership and development of macro- (plot a) and microvascular (plot b) complications of type 2 diabetes after adjustment for hypertension, statin use and serum cholesterol. Clusters are defined as Clinically Undifferentiated High Polygenic Susceptibility Diabetes (CUPS), Mild Age-Related Diabetes (MARD), Severe Insulin-Resistant Diabetes (SIRD), Insulin-Resistant Diabetes (IRD) and Mild Obesity-Related Diabetes (MOD).

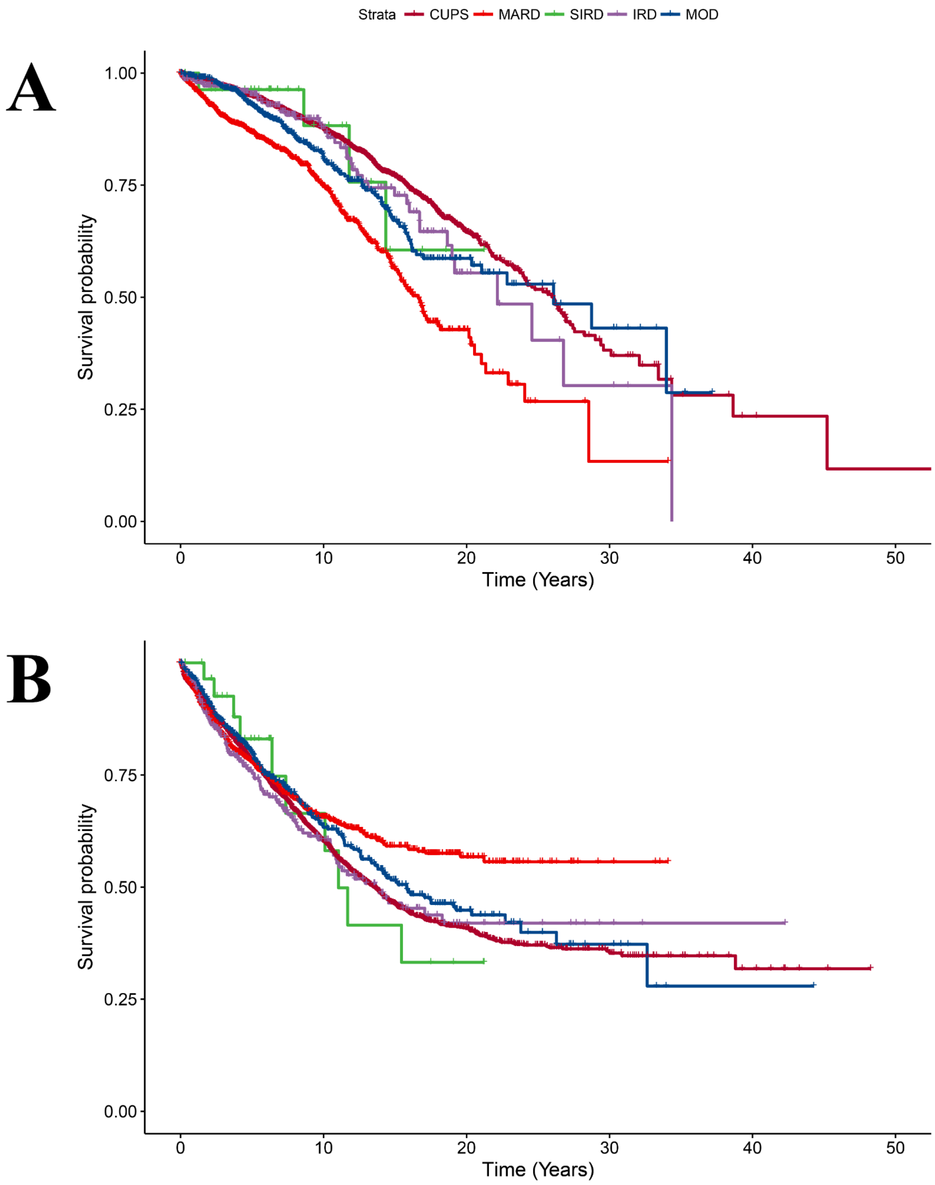
