## Supplementary File 1 for "Harnessing the power of polygenic risk scores to predict type 2 diabetes and its subtypes in a high-risk population of British Pakistanis and Bangladeshis in a routine healthcare setting"

*Quality control and imputation of genotype data in Genes & Health*

Standard quality control of samples (N=22,944) and variants was performed in Illumina's GenomeStudio software. For duplicate samples and variants, the one with the highest call rate was retained. Variants with call rate <0.99, minor allele frequency (MAF) <1% were removed using plink v1.9. Variants that failed the Hardy-Weinberg test (P-value $<1\times10^{-6}$) in 64% of the samples with low levels of autozygosity (i.e. proportion of the genome in runs of homozygosity <0.5%) were excluded.

To identify individuals of British Bangaldeshi and British Pakistani ancestry, we merged G&H with individuals of South Asian ancestry from the Human Genome Diversity Project (HGDP) [1] and 1000 Genomes Project Phase 3 [2]. We performed principal component analysis (PCA) in HGDP and 1000 Genomes samples, and projected G&H samples onto the same PC space using EIGENSOFT [3]. We then performed the uniform manifold approximation and projection dimension reduction method (UMAP) [4] on the top 10 PCs using the R package "uwot". We assigned British Pakistani and Bangladeshi ancestry to G&H samples that were in the same cluster with samples of Pakistani and Bangladeshi ancestry from external cohorts, respectively. We excluded individuals with reported ethnicity inconsistent with the genetically inferred ancestry.

The Michigan Imputation Server was used to perform imputation with the Genome Asia pilot reference panel. Palindromic SNPs and SNPs that were not found in the reference panel (including those with mismatched alleles) were excluded before imputation. We used different sets of imputed SNPs for different analyses in G&H:

- SNPs with INFO score ≥0.3 and MAF ≥0.1% (N=9,527,863) for constructing PRS.
- SNPs with INFO ≥0.7 (and MAF ≥0.5% for genome-wide association analysis (GWAS)
- SNPs with INFO>0.7 and MAF>1% for genetic correlation and colocalisation.

*Transferability of previously identified GWAS loci*

Previously identified loci associated with type 2 diabetes were obtained from the European-ancestry (EUR) GWAS (N= ~1.1 million) by Vujkovic *et al.* [5]. Unlike the construction of PRS for which the multi-ancestry GWAS data were used to increase power, we used European-ancestry GWAS to assess transferability of GWAS loci. The list of lead variants at each locus were from Supplementary Table 6 in the Vujkovic *et al* study. We performed GWAS of type 2 diabetes in G&H with SAIGE[6] and adjusted for sex, age, $age^2$, and first 20 genetic PCs. We further excluded individuals with an EHR clinical code associated with being "at risk" of type 2 diabetes (**Table S1**), but not having developed the condition, leaving 12,785 individuals in GWAS. Following Huang *et al.* [7], we assessed whether previously identified loci were reproducible in G&H at P-value <0.05. Credible sets for established loci consisted of lead (independent) variant and proxy SNPs ($r^2$ >=0.8) within a 50kb window (based on the 1000 Genomes EUR data) of the sentinel variant and with p-value $<100\times p_{sentinel}$. We assessed 338 loci from the Vujkovic *et al.* study that had variants in credible sets well-imputed in G&H. Expected power for replication was estimated accounting for the original effect size, the allele frequency and sample size in G&H. The number of loci that were expected to be transferable was estimated by summing up the power of lead variants across identified loci. We defined 'transferable loci' as those with at least one variant in the credible set associated with type 2 diabetes (P-value <0.05) in the same direction in G&H. We observed 76 (22.5%) transferable loci and the ratio of the number of observed transferable loci to that of expected (28.1%) was 0.80. The high transferability was consistent with other cardiometabolic traits (Observed/Expected = 0.62 for coronary artery disease and 1.0–1.2 for BMI, lipids, and blood pressure) that were reported in the same cohort [7]. We defined 'non-transferable' loci as those with ≥1 variant in the credible set with >80% power but that had no variant in the credible set significant at P-value <0.05 and no variant within the 50kb window significant at P-value $<1\times10^{-3}$. We did not observe any loci that were well powered but not transferable in G&H.

Trans-ancestry colocalisation analysis was performed using TEColoc to assess whether a transferable locus shared the same causal variant between BPB and UK Biobank (UKBB) EUR populations [8]. For each transferable locus, we used variants in a 50kb window that existed in both cohorts, and assessed loci with the proportion of overlapping SNPs ≥10% in both cohorts. We used a significance threshold of P-value <0.05 to determine evidence a causal variant was shared. We assessed 27 transferable loci and 9 (33.3%) of them showed strong evidence of shared causal variants. The proportion of transferable loci with shared causal variants for type 2 diabetes was lower than blood lipids (47-61%) and slightly higher than BMI (26%), which have also been investigated in G&H[7].

We applied the Popcorn algorithm to estimate the trans-ancestry genetic correlation between BPB and UKBB EUR populations using GWAS summary statistics [9]. LD scores were estimated using 1000 Genomes SAS and EUR populations, excluding the major histocompatibility complex region.

*Prediction of incident type 2 diabetes.*

*QDiabetes Score Estimation*: For each individual, QDiabetes model A, B and C scores were estimated using the R package "QDiabetes" [10]. Model A provides estimates based on clinical data from the EHR including age, ethnicity, family history, comorbidities, and prescribed medications. Model B, the model with highest predictive value, is calculated using the same variables as model A, plus fasting plasma glucose (FPG), while model C is composed of model A plus HbA1c. Due to fewer participants without an established diagnosis of type 2 diabetes having recorded FPG or HbA1c, numbers available for analysis were lower for model B (n = 4,334) and model C (n = 864) than model A (n = 13,648). We applied multiple imputation to fill in missing data for BMI and the Townsend Deprivation Index using the R package "MICE" [11].

*Integrated Risk Tool (IRT) Score Estimation*: There was no correlation between PRS and QDiabetes scores (Pearson's coefficients -0.03, 0.08, 0.13 for QDiabetes models A, B, and C respectively; associated p-values 0.31, 0.18, 0.16). In brief, the QDiabetes score was converted to an odds ratio, and multiplied by the odds calculated from the individual's PRS given their QDiabetes score, calculated from a logistic regression model incorporating an interaction effect between QDiabetes and PRS. This logistic regression model was separately trained on male and female participants.

*Assessment of IRT Performance:* The net reclassification index (NRI) was calculated as the sum of (1) the proportion of individuals who subsequently developed type 2 diabetes correctly reclassified as high risk, minus the proportion of individuals who subsequently developed type 2 diabetes incorrectly reclassified as low risk; and (2) the proportion of individuals who did not develop type 2 diabetes correctly reclassified as low risk, minus the proportion of individuals who did not develop type 2 diabetes incorrectly reclassified as high risk. We chose 10% as the threshold for classification of high and low risk individuals, in line with previously published literature. NRI performance was assessed in the entire analysed sample, plus in age-by-sex subgroups, with 40 years chosen as the threshold for high/low age group comparison to reflect (1) a mean age at type 2 diabetes diagnosis of 49.1 years and median of 47.8 years in our cohort, (2) a mean age at study entry of 39.1 years, and (3) the fact that routine NHS health checks that include type 2 diabetes screening are offered at age 40. NRI confidence intervals were calculated using bootstrapping (number of iterations = 1,000). The characteristics of reclassified individuals were compared with descriptive statistics. C-Indices were calculated from Cox proportional hazard models run using the R package "survival".

*Prediction of transition from gestational diabetes mellitus (GDM) to type 2 diabetes:* Characteristics of women with a history of GDM who did and did not develop type 2 diabetes were compared in multivariate logistic regression models that included risk factors used to construct the QDiabetes score. The association between PRS and type 2 diabetes in women with a history of GDM was assessed in Cox proportional hazard models controlling for (1) QDiabetes score or (2) unadjusted clinical risk factors used to construct the QDiabetes score

1. Li, J. Z. *et al.* Worldwide human relationships inferred from genome-wide patterns of variation. *Science* **319**, 1100–1104 (2008).
2. 1000 Genomes Project Consortium *et al.* A global reference for human genetic variation. *Nature* **526**, 68–74 (2015).
3. Patterson, N., Price, A. L. & Reich, D. Population structure and eigenanalysis. *PLoS Genet.* **2**, e190 (2006).
4. McInnes, L., Healy, J. & Melville, J. UMAP: Uniform Manifold Approximation and Projection for Dimension Reduction. *arXiv [stat.ML]* (2018).
5. Vujkovic, M. *et al.* Discovery of 318 new risk loci for type 2 diabetes and related vascular outcomes among 1.4 million participants in a multi-ancestry meta-analysis. *Nat. Genet.* **52**, 680–691 (2020).
6. Zhou, W. *et al.* Efficiently controlling for case-control imbalance and sample relatedness in large-scale genetic association studies. *Nat. Genet.* **50**, 1335–1341 (2018).
7. Huang, Q. Q. *et al.* Transferability of genetic loci and polygenic scores for cardiometabolic traits in British Pakistanis and Bangladeshis. *medRxiv* 2021.06.22.21259323 (2021).
8. Kuchenbaecker, K. *et al.* The transferability of lipid loci across African, Asian and European cohorts. *Nat. Commun.* **10**, 4330 (2019).
9. Brown, B. C., Asian Genetic Epidemiology Network Type 2 Diabetes Consortium, Ye, C. J., Price, A. L. & Zaitlen, N. Transethnic Genetic-Correlation Estimates from Summary Statistics. *Am. J. Hum. Genet.* **99**, 76 (2016).
10. Feakins, B. G. Type 2 Diabetes Risk Calculator [R package QDiabetes version 1.0-2]. (2021).
11. van Buuren, S. & Groothuis-Oudshoorn, K. mice: Multivariate Imputation by Chained Equations in R. *J. Stat. Softw.* **45**, 1–67 (2011).
